# The effect of reopening policy on COVID-19 related cases and deaths

**DOI:** 10.1101/2020.06.25.20139840

**Authors:** Qiyao Zhou

## Abstract

By May 29, 2020, all 50 states in the United States had reopened their economies to some extent after the coronavirus lockdown. Although there are many debates about whether states reopened their economies too early, no study has examined this effect quantitatively. This paper takes advantage of the daily cases, deaths, and test data at the state level, and uses the synthetic control method to address this question. I find that reopening the economy caused an additional 2000 deaths in the 6 states (Alabama, Colorado, Georgia, Mississippi, Tennessee, and Texas) that reopened before May 1^st^ by three weeks after reopening. It also increased daily confirmed cases by 40%, 52%, and 53% after the first, second, and third week of reopening, respectively. Moreover, contrary to scientists’ prescription that expanding tests is a necessary condition for reopening, these states witnessed a decline in daily tests by 17%, 47%, and 31% after the first, second, and third week of reopening, respectively.

## 1. Introduction

> *If states reopen prematurely and don’t have the capability to handle new cases, “the consequences could be really serious” and more outbreaks can occur*.
>
> -Anthony Fauci, May 12 2020

The outbreak of coronavirus in 2020 has become one of the most serious challenges to public health not only in the United States but also throughout the world. In response to the rapid spread of the coronavirus, 43 states and Washington DC issued the stay-at-home orders and shut down their economies. On one hand, the stay-at-home order is believed to increase the fraction of people who stay at home all day (Guo, 2020), reduce COIVD-19 cases and save lives (Fowler et al., 2020; Friedson et al., 2020; Lyu and Wehby, 2020). On the other hand, the economic cost of stay-at-home order was large, and increased unemployment significantly. On April 24^th^, Alaska was the first to reopen its economy, followed by Colorado (reopened on April 26^th^), Montana (reopened on April 26^th^), and Mississippi (April 27^th^), etc.

There are fierce policy debates about whether some states in the United States reopened their economies too fast. Stock (2020) asked the following questions: *“What are the consequences of reopening now, as opposed to* (*say*) *waiting until deaths decline further? Among NPIs that have similar effects on the paths of infections and deaths, are some more economically desirable than others? How can one most effectively reopen the economy while achieving some public health objective, whether flattening the curve or sharply reducing infections and deaths?”*

Thus far, the answers to these questions are limited. Alvarez et al. (2020) constructed a planner’s dynamic planning problem, and found that the optimal policy prescribes a lockdown for around two weeks, followed by a gradual loosening of restrictions in the following four months. Rampini (2020) calibrated an SIR model with heterogeneous population and suggested a sequential approach to reopening the economy. One consensus that current research has reached is that sufficient testing is a necessary condition to reopen the economy (Taipale et al., 2020; Alvarez et al., 2020; Berger et al., 2020).

The goal of this study is to quantify the effect of reopening the economy on daily confirmed cases and deaths. I use the synthetic control method (Abadie and Gardeazabal, 2003; Abadie et al., 2010) to estimate the effect of this policy in the states that were the first to open. The main advantage of this approach is that it requires fewer observations and imposes fewer functional form restrictions to construct a robust estimator compared with the difference-in-difference method.

Using the state-level daily confirmed COVID-19 cases, COVID-related deaths data, and test data from March 1^st^ to May 24^th^, I comprehensively analyze the effect of reopening policy. George Floyd’s death on May 25^th^ sparked nationwide protests in subsequent weeks. These widespread protests may be linked to the spread of the coronavirus and may bias the analysis of policies to reopen the economy if I include the data after May 25^th^.

My preferred results suggest that reopening policies causes an additional 2000 deaths in the 6 states that reopened before May 1^st^ by three weeks after reopening. Reopening also significantly increased the daily confirmed cases in these 6 states by more than 1000. However, the increase in the cases was not due to the increase in tests conducted. On the contrary, the number of tests conducted in these states significantly decreased. All these results suggest that reopening the states too early incurred substantial public health costs.

The COVID-19 outbreak has been accompanied by a cascade of academic research. Many papers discuss the effect of social distancing on flattening the curve (Kapoor et al., 2020; Mangrum and Niekamp, 2020). However, there is little evidence on the impact of reopening the economy. My study mainly focuses on the costs of reopening the economy. The results of my paper shed light on its potential consequences, and lays the foundations to answering questions asked by Stock (2020): when and how to reopen the economy?

Section 2 introduces the data and empirical methods I use to study the impact of reopening the state on confirmed cases, deaths, and tests. Section 3 discusses the results. A conclusion is drawn in section 4.

## 2. Methods

### 2.1 Data

State-level data on coronavirus daily confirmed cases, deaths comes from Killeen et al. (2020)’s JHU dataset. The coronavirus test data come from the New York Times dataset and is aggregated by the *“covidtracking website”*^*2*^. The CDC in some states conflated viral and antibody tests at the beginning of the coronavirus outbreak, and reported both. Then they adjusted this data and reported the viral test results only. Therefore, the number of tests in some states might become negative at the date when they adjusted the test data. It is worthwhile to note that the test data are more volatile and of lower quality compared with confirmed cases and deaths data. I collect the state-level confirmed COVID-19 cases, COVID-19 related deaths, and number of tests data from March 1st to May 24^th^ on a daily basis.

The date the state reopened the economy comes from the New York Times dataset, and the Kaiser Family Foundation ^3^. The treatment group includes the states that reopened the economy before May 1^st^, the remaining states are assigned to the control group. I also restrict our control group to the states that reopened after May 10^th^ in Appendix 2, and the results are still robust. Table 1 lists the states that reopened before May 10^th^.

**Table 1.** The reopening date of states

| State | reopening date | state | reopening date |
| --- | --- | --- | --- |
| Alaska | 2020/4/24 | Missouri | 2020/5/3 |
| Colorado | 2020/4/26 | Kansas | 2020/5/3 |
| Montana | 2020/4/26 | Indiana | 2020/5/4 |
| Mississippi | 2020/4/27 | South Carolina | 2020/5/4 |
| Texas | 2020/4/30 | Florida | 2020/5/4 |
| Georgia | 2020/4/30 | North Carolina | 2020/5/8 |
| Tennessee | 2020/4/30 | Pennsylvania | 2020/5/8 |
| Idaho | 2020/4/30 | Rhode Island | 2020/5/8 |
| Alabama | 2020/4/30 |  |  |

Source: New York Times dataset and Kaiser Family Foundation.

### 2.2 Estimation

I use the synthetic control method introduced by Abadie and Gardeazabal (2003), and Abadie et al. (2011) to estimate the effect of reopening the economy on the daily confirmed cases and daily COVID-19 related deaths in the nine states that reopened before May 1^st^. The synthetic control method allows more flexible functional forms and imposes fewer restrictions on the observations.

The estimation procedure can be summarized as follows:

In the first step, I choose the optimal weight W^*^ to minimize:

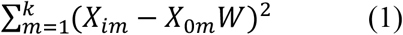

Where *X*_*im*_ is the value of the m^th^ variable for the 9 treated states. *X*_0*m*_ is a 1 × n vector containing the values of the m^th^ variable for the units in our donor pool, which includes n=42 states (and the District of Columbia) that reopened or planned to reopen after May 1^st^. As noted above, I also consider as an alternate control group the states that reopened or planned to reopen after May 10^th^, and the results remain robust.

In the second step, the effect of reopening the economy can be estimated as:

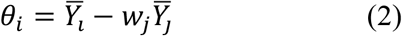

Where *Y*_*i*_ and *Y*_*j*_ are the outcome variables (i.e., daily confirmed cases, deaths, and the number of tests).

In step 2, I use the bootstrapping method to conduct hypothesis testing. The estimated effect 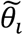 is denoted by:

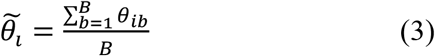

The estimated variance is:

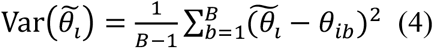

where B denotes the total bootstrapping times. B=50 in this study. θ_*ib*_ denotes the synthetic control estimator for state i at time b.

The observables I choose to match across the treatment and control pools include the outcome variables (i.e., daily confirmed cases, deaths, and number of tests), and a set of controls that includes population density and hospital beds per capita. I report the results that use only the outcomes variables as the matching criterion. The results remain unchanged if I add a set of controls.

With regard to the time window, I largely follow (Friedson et al., 2020). My primary strategy is to generate a synthetic control that closely approximates coronavirus cases (and deaths, and tests) in the treated states in each of seven pre-treatment days. When I extend the pre-treatment days to 14 in Appendix 3, the results remain quite robust.

One drawback of the synthetic control method is that the variable of interest (i.e., daily confirmed cases, deaths, and number of tests) in the treated states should be written as a convex combination of the same variables in the donor pool. Therefore, it is hard to find an ideal synthetic state for some states with the highest daily confirmed cases or deaths, and for some states with the lowest daily confirmed cases and deaths (e.g., Alaska, Montana, and Idaho). I delete Alaska, Montana, and Idaho from the treated pool, and analyze the effect of reopening the economy in the 6 remaining states (Alabama, Colorado, Georgia, Mississippi, Tennessee, and Texas) that reopened before May 1^st^.

## 3. Results

In this section, I present the synthetic control results using the daily confirmed cases, COVID-19 related deaths, and number of tests conducted as the variable of interest. In Appendix 1, I present parallel results that uses the same variable of interest but on a per million population basis. It is quite similar to our main results.

### 3.1 Reopening the economy and daily confirmed cases

Figure 1 compares daily confirmed cases for each of the 6 states that reopened before May 1^st^ and their synthetic control states. I assign weights to synthetic control states based on the daily confirmed cases on each of 7 pre-treatment dates. The y-axis is the daily confirmed case in thousands, and the x-axis is the date. The dashed line in the middle represents the effective date when the state reopened the economy.

**Figure 1.**
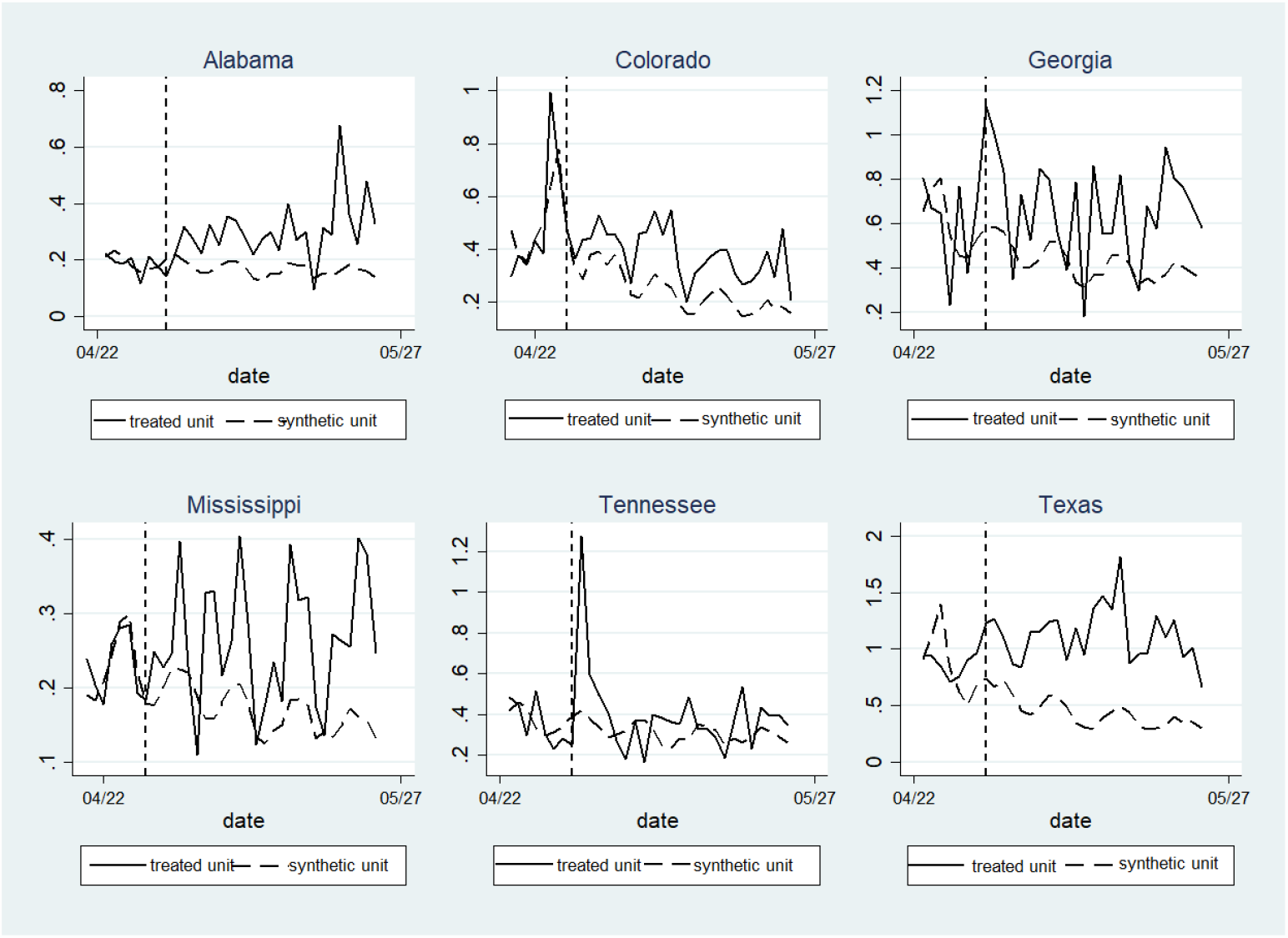
Synthetic control results for daily confirmed cases.

Trends in daily confirmed cases for the treated states before they reopened the economy do not show a clear declining pattern. The results in Figure 1 suggest that the synthetic control states match quite well most of the treated state during the pre-treatment periods (although Texas is not matched very well). However, after the treated states reopened the economy, two lines diverge markedly, with the daily confirmed cases in treated states that reopened the economy before May 1^st^ increasing much faster than the synthetic control states.

I then check the timing effect of the reopening policy and conduct the statistical inference using bootstrapping methods. The results are reported in Table 2. Note that in Table 2. Week 0 refers to the week before the reopening policy. Weeks 1, 2, and 3 refer to the weeks after the reopening policy, respectively. The column treated-synthetic refers to the difference in terms of the daily confirmed case between the treated state and the synthetic state. The coefficients reported in Table 2 could be interpreted as the difference in terms of the number of daily COVID-19 confirmed cases between the treated states and the synthetic control states at week i. For instance, 74.3 at column 3 row 3 means that on average Colorado has 74.3 more COVID-19 confirmed cases on a daily basis compared with the synthetic control states for Colorado.

**Table 2.** Estimated effect of reopening on daily confirmed cases

| week | state | treated-synthetic | std | week | state | treated-synthetic | std |
| --- | --- | --- | --- | --- | --- | --- | --- |
| 0 | Colorado | -2.6 | 1.62 | 0 | Mississippi | 0.4 | 1.82 |
| 1 | Colorado | 74.3 | 48.72 | 1 | Mississippi | 30.4* | 15.98 |
| 2 | Colorado | 175.4** | 74.14 | 2 | Mississippi | 102.6*** | 19.93 |
| 3 | Colorado | 122.3* | 64.64 | 3 | Mississippi | 98.8*** | 15.61 |
| 0 | Texas | -1.8 | 2.12 | 0 | Georgia | -0.3 | 2.14 |
| 1 | Texas | 489.3*** | 65.77 | 1 | Georgia | 273.4*** | 26.18 |
| 2 | Texas | 754.7*** | 72.79 | 2 | Georgia | 175.5*** | 28.30 |
| 3 | Texas | 807.8*** | 72.30 | 3 | Georgia | 217.7*** | 29.44 |
| 0 | Tennessee | -1.4 | 2.60 | 0 | Alabama | -1.0 | 2.23 |
| 1 | Tennessee | 149.6*** | 8.20 | 1 | Alabama | 67.9*** | 3.69 |
| 2 | Tennessee | 58.9*** | 8.65 | 2 | Alabama | 125.7*** | 4.08 |
| 3 | Tennessee | 25.1*** | 8.91 | 3 | Alabama | 172.1*** | 4.17 |

The results in Table 2 support the patterns reported in Figure 1. I find that there doesn’t exist a significant difference in daily confirmed cases between treated and synthetic control states before the date when the treated states reopened the economy. However, this difference becomes large and significant after the reopening policy. And the gap reaches its peak 2 weeks after reopening, then remains relatively constant in the 3^rd^ week. Using the average daily confirmed cases during the pre-treatment week as the benchmark, I find that the daily confirmed cases increase by 40% in the first week after reopening the economy, and this number increases to 52% and 53% in the second and the third weeks after the policy, respectively. The analysis suggests that if the goal of the policy-maker is to reduce the transmission of the coronavirus, then it may have not been successful for these 6 states.

Note: *** p<0.01, ** p<0.05, * p<0.1. Standard errors are estimated by bootstrapping 50 times.

### 3.2 Reopening the economy and deaths

I then change the variable of interest to daily COVID-19 related deaths and conduct a similar analysis as in section 3.1. Figure 2 reports the synthetic control results for daily deaths. The y-axis in Figure 2 represents daily deaths, and the x-axis denotes the date. I assign weights to synthetic control states based on the daily deaths on each 7 pre-treatment dates.

**Figure 2.**
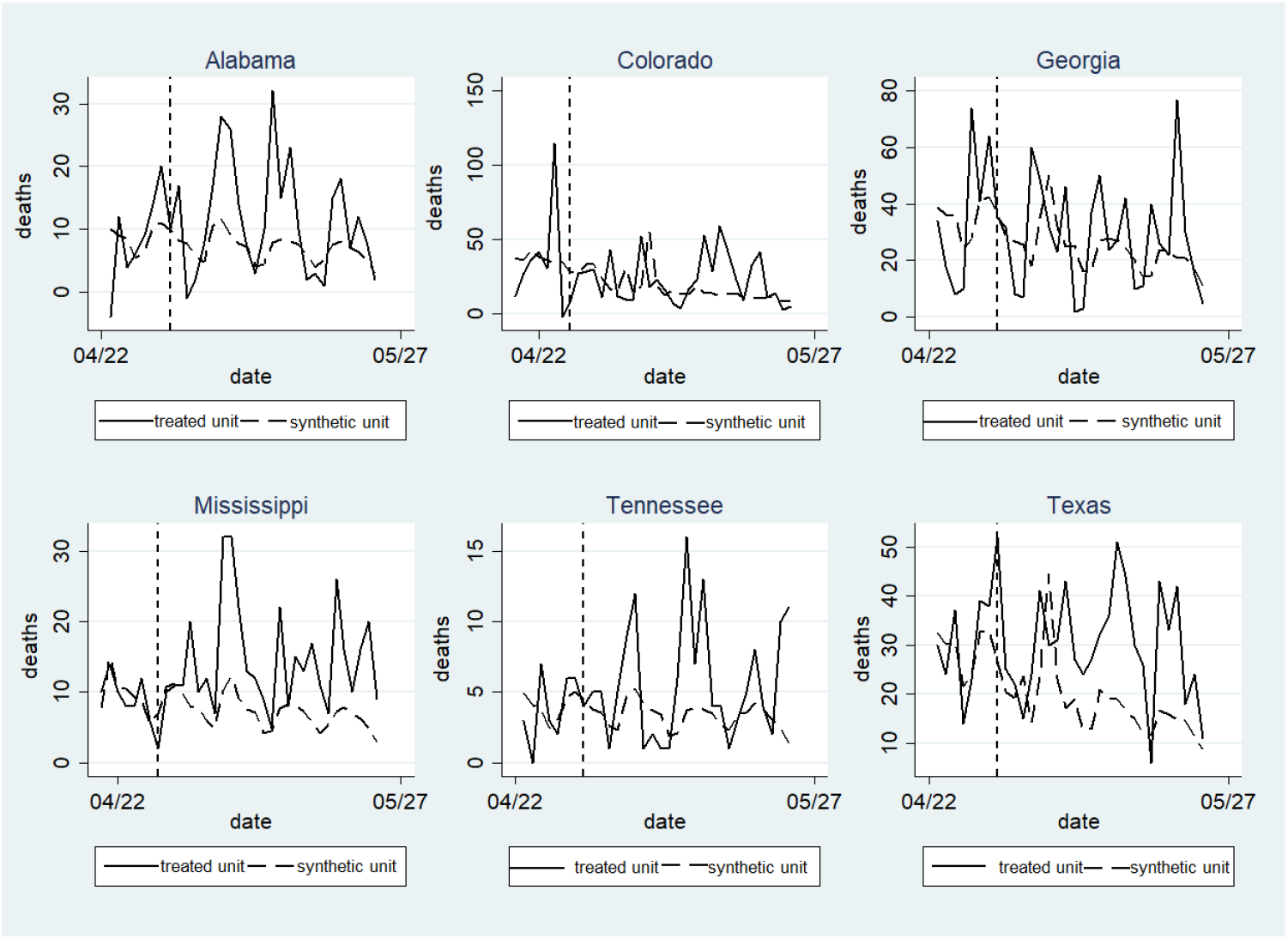
Synthetic control results for daily deaths.

I find that the daily COVID-19 related deaths are more volatile and harder to predict than the daily confirmed cases, especially for Georgia and Colorado. Nonetheless, I still find that daily deaths between the treated states and the synthetic states start to diverge after the treated states reopened the economy. The results in Figure 2 suggest that reopening the state too early greatly increases the deaths.

I check the timing effect of the reopening policy and conduct statistical inference using the bootstrapping method. The results are reported in Table 3. The notation in Table 3 remains exactly the same as in Table 2, with week 0 denoting the pre-treatment periods, and week 1, 2 and 3 denoting the first, second and the third week after reopening the economy. The coefficients reported in Table 3 can be interpreted as the difference in the number of daily COVID-19 related deaths between the treated states and the synthetic control states at week i. For instance, 2.6 in column 3 row 3 means that on average Colorado has 2.6 more COVID-19 related deaths on a daily basis compared with its synthetic controls.

**Table 3.** Estimated effect of reopening on daily deaths

| week | state | treated-synthetic | std | week | state | treated-synthetic | std |
| --- | --- | --- | --- | --- | --- | --- | --- |
| 0 | Colorado | -0.0 | 0.12 | 0 | Mississippi | 0.0 | 0.11 |
| 1 | Colorado | -2.6*** | 0.43 | 1 | Mississippi | 2.1*** | 0.19 |
| 2 | Colorado | -4.46*** | 0.67 | 2 | Mississippi | 10.2*** | 0.21 |
| 3 | Colorado | 18.2*** | 0.17 | 3 | Mississippi | 6.4*** | 0.21 |
| 0 | Texas | 0.12 | 0.15 | 0 | Georgia | 0.10 | 0.17 |
| 1 | Texas | 5.2** | 0.28 | 1 | Georgia | 0.24 | 0.41 |
| 2 | Texas | 13.6** | 0.51 | 2 | Georgia | 2.1*** | 0.46 |
| 3 | Texas | 18.0** | 0.47 | 3 | Georgia | 4.2*** | 0.44 |
| 0 | Tennessee | -0.1** | 0.05 | 0 | Alabama | -0.04 | 0.10 |
| 1 | Tennessee | 2.05*** | 0.05 | 1 | Alabama | 3.2*** | 0.10 |
| 2 | Tennessee | 1.59*** | 0.05 | 2 | Alabama | 8.2*** | 0.12 |
| 3 | Tennessee | 2.09*** | 0.07 | 3 | Alabama | 3.6*** | 0.11 |

Note: \*\*\* $p < 0.01$ , \*\* $p < 0.05$ , \* $p < 0.1$ . Standard errors are estimated by bootstrapping 50 times.

The results in Table 3 support the patterns reported in Figure 2. The results suggest that the difference in daily deaths between treated and synthetic control states is negligible during the pre-treatment period. Then daily deaths in treated states increase by around 9% compared with the synthetic control states in the first week after reopening the economy. This number increases to 25%, and 38% in the second and the third week after reopening the economy, respectively. I find that there is a one to two week lag in the increase in deaths compared with the increase in daily confirmed cases. This result is consistent with Wang et al., (2020) and Wang et al., (2020) who argue that the median time from symptom onset to death is around 2 weeks. Converting the percentage to numbers, the above results suggest that reopening the economy early causes an additional of 2000 deaths in these 6 states within 3 weeks after the policy.

### 3.3 Reopening the economy and the number of tests

One consensus that scientists have reached is that expanding testing capacity is a necessary condition for reopening the economy. Testing aggressively could discover new cases at the early stage, and permit efficient contact tracing. Taipale et al. (2020) propose testing every individual repeatedly, and then isolating the infected individuals. Peto et al. (2020) argued that aggressively testing is the lockdown exit strategy for the UK. They estimated that for a local population of 200,000 people, with 90% compliance, this will require 26,000 tests per day.

In this section, I check whether states expanded their test capacity after they reopened the economy. If the daily tests performed significantly expanded in these states, the increase of daily confirmed cases does not necessarily imply that the incidence of the coronavirus per million people increased.

Figure 3 reports the synthetic control results for the number of tests performed^4^, with the same notation as in Figure 1 and 2. I assign weights to synthetic control states based on the number of daily tests on each of 7 pre-treatment dates. Note that because of the adjustment of the conflating of the viral and antibody test data in Mississippi, the number of tests performed on May 23 becomes negative. Although the test data is more volatile and of lower quality compared with confirmed cases and deaths data, it could still provide some suggestive evidence as to how tests were conducted after the reopening of the economy in these states.

**Figure 3.**
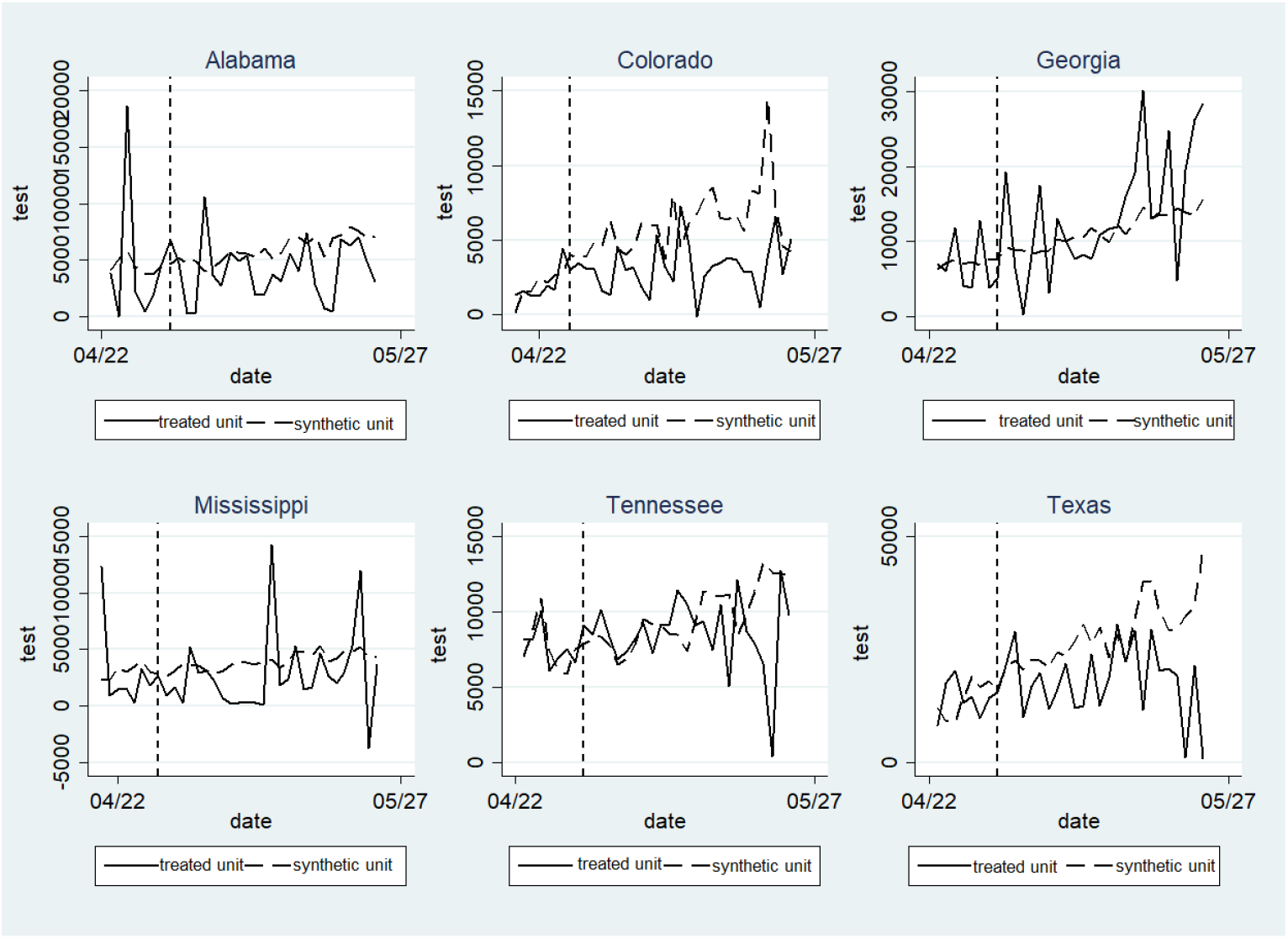
Synthetic control results for daily tests.

The results in Figure 3 suggest that except for Georgia, the number of tests performed in the treated states after they reopened the economy decreased compared with their control counterparts.

I conduct hypothesis testing and check the timing effect of reopening the economy on the number of tests. The results are reported in Table 4, using the same notation as in Table 2 and Table 3. The coefficients in Table 4 can be interpreted as the difference in terms of the number of tests performed on a daily basis between the treated states and the synthetic control states at week i. For instance, −1603.4 at column 3 row 3 means that on average Colorado conducted 1603.4 fewer tests on a daily basis compared with the synthetic control states for Colorado.

**Table 4.**
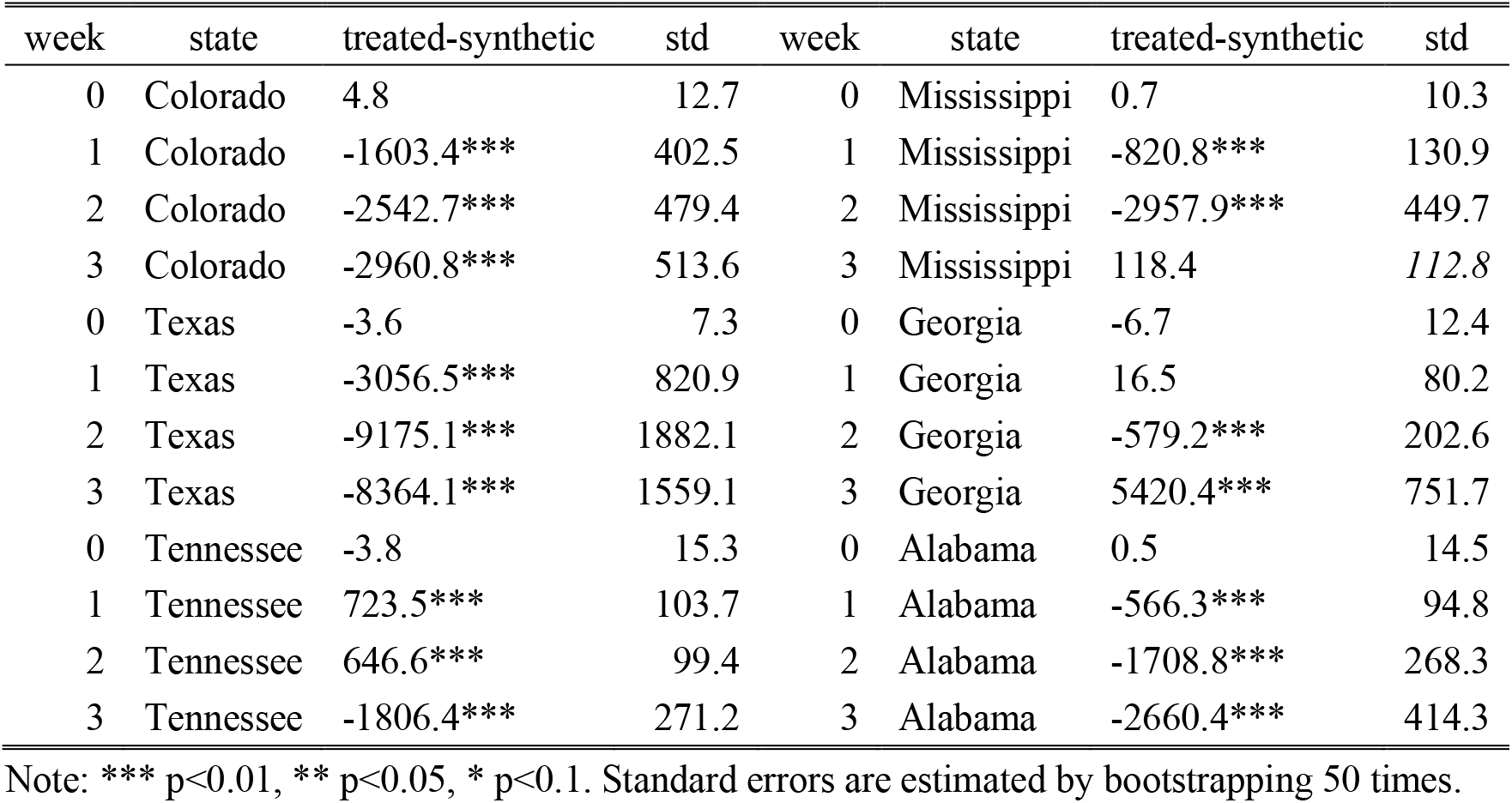
Estimated effect of reopening on daily tests

| week | state | treated-synthetic | std | week | state | treated-synthetic | std |
| --- | --- | --- | --- | --- | --- | --- | --- |
| 0 | Colorado | 4.8 | 12.7 | 0 | Mississippi | 0.7 | 10.3 |
| 1 | Colorado | -1603.4*** | 402.5 | 1 | Mississippi | -820.8*** | 130.9 |
| 2 | Colorado | -2542.7*** | 479.4 | 2 | Mississippi | -2957.9*** | 449.7 |
| 3 | Colorado | -2960.8*** | 513.6 | 3 | Mississippi | 118.4 | 112.8 |
| 0 | Texas | -3.6 | 7.3 | 0 | Georgia | -6.7 | 12.4 |
| 1 | Texas | -3056.5*** | 820.9 | 1 | Georgia | 16.5 | 80.2 |
| 2 | Texas | -9175.1*** | 1882.1 | 2 | Georgia | -579.2*** | 202.6 |
| 3 | Texas | -8364.1*** | 1559.1 | 3 | Georgia | 5420.4*** | 751.7 |
| 0 | Tennessee | -3.8 | 15.3 | 0 | Alabama | 0.5 | 14.5 |
| 1 | Tennessee | 723.5*** | 103.7 | 1 | Alabama | -566.3*** | 94.8 |
| 2 | Tennessee | 646.6*** | 99.4 | 2 | Alabama | -1708.8*** | 268.3 |
| 3 | Tennessee | -1806.4*** | 271.2 | 3 | Alabama | -2660.4*** | 414.3 |

Note: \*\*\* $p < 0.01$ , \*\* $p < 0.05$ , \* $p < 0.1$ . Standard errors are estimated by bootstrapping 50 times.

The results in Table 4 support the pattern in Figure 3. The difference in the number of tests performed on a daily basis is negligible between the treated and synthetic control states during the pre-treatment period (week 0). However, except for Georgia, the number of tests performed in the treated states significantly decreases after the reopening of the economy. Using the number of tests performed in treated states at week 0 (pre-treatment week) as a benchmark,^5^ the number of tests performed in the treated states decreases by around 17% in the first week after they reopened. This number becomes 47%, and 31% in the second and the third weeks, respectively.

The analysis underscores two points. First, the increase in the number of daily confirmed cases in the states that reopened before May 1^st^ doesn’t come from the expansion of the tests. On the contrary, the test number shrinks after reopening the economy. Second, these six states are not well-prepared for reopening. Lacking tests and reopening the economy too early is accompanied by significant public health costs.

Appendix 1 contains parallel results on a per million people basis. I find that reopening the states too early for the six states increased their daily confirmed cases on a per million basis by 155, 200, and 207 after the first, second, and third week of reopening, respectively, with Texas witnessing the highest increase. It increased the daily deaths on a per million basis by 1.5, 4.4, and 7.6 after the first, second, and third week of reopening, respectively, with Texas, Alabama, and Mississippi witnessing the highest increase. It decreased the tests on a per million basis by 978, 2611, and 1731 after the first, second, and third week of reopening, respectively.

## 4. Conclusions

By May 29, 2020, all 50 states in the United States had reopened their economies to some extent after the coronavirus lockdown. There have been many policy debates about when a state should reopen its economy. On one hand, reopening the economy reduces unemployment. On the other hand, reopening the economy prematurely may increase the spread of the coronavirus. There have been few rigorous, quantitative studies of this topic.

This study uses the synthetic control method to rigorously study the short-term effect of reopening the economy on the daily confirmed COVID-19 cases, daily deaths, and daily tests in 6 states (Alabama, Colorado, Georgia, Mississippi, Tennessee, and Texas) that reopened before May 1^st^. I find that all these states experienced a sharp increase in both confirmed COVID-19 cases, and COVID-19 related deaths. Except for Georgia, the other 5 states saw a decline in the number of tests performed after they reopened the economy. The magnitude of the increase in confirmed cases and deaths is substantial. It resulted in an additional 2000 deaths in the 6 states compared with their control counterparts. It also increased daily confirmed cases by 40%, 52%, and 53% after the first, second, and third weeks of reopening, respectively. However, the increase of confirmed COVID-19 cases and deaths was not due to the expansion of tests. On the contrary, the number of tests performed on a daily basis decreased by 17%, 47%, and 31% after the first, second, and third weeks of reopening, respectively. The findings are robust to the pre-treatment time windows and the choice of observables with which the donor states are weighted.

My study provides some evidence that 6 states reopened prematurely: reopening increased both deaths and transmission of the virus. There are, however, additional questions that future research must address. First, my work estimates the public health costs associated with reopening the economy. Future work needs to focus on the economic benefits of reopening. Second, it is critical to study what is the most efficient way to reopen the economy from a public health perspective. If expanding tests are the best answer to this question, then how many tests are needed to monitor the transmission of COVID-19 after reopening the economy? Third, this study focuses on the short-term effects of early reopening of the economy early: What are the long-term effects?

## Data Availability

Data and STATA do files are available on request.

## Acknowledgements

I am indebted to Maureen Cropper for her constructive suggestions and encouragement during this project. I thank Ziyang Chen, and Jiangnan Zeng for valuable comments.

## Appendix 1

In this Appendix I change the variables of interest from daily COVID-19 confirmed cases, daily COVID-19 related deaths, and daily tests performed to daily COVID-19 confirmed cases per million population, daily COVID-19 related deaths per million population, and daily tests performed per million population, respectively. The results are reported in Tables A1-A3, using the same notation as in Tables 2-4.

The results in Table A1 are similar to the results in Table 2: reopening the economy in the 6 states significantly increases the daily confirmed cases per million population. Texas saw the largest increase, with daily confirmed cases increasing by 489.3 per million population in the first week after reopening the economy compared with its control counterparts. This number increased to 745.7 per million population, and 807.8 per million population in the second and the third weeks after reopening respectively.

**Table A1.**
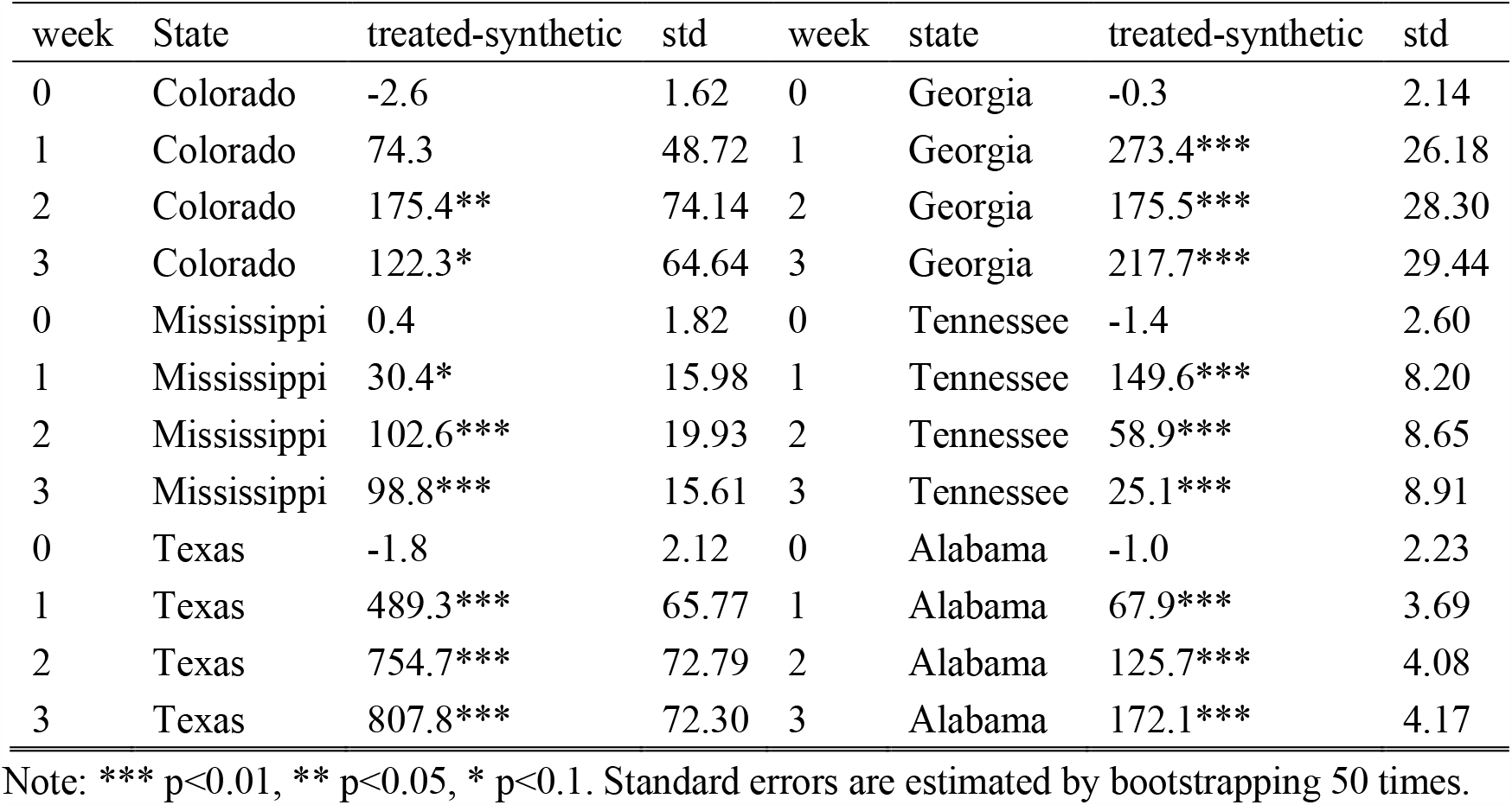
Estimated effect of reopening on daily confirmed cases per million

The results reported in Table A2 support the findings in Table 3: Reopening the economy in the six states significantly increased the daily COVID-19 related deaths per million population. Texas also saw the largest increase in COVID-19 related deaths per million population.

**Table A2.** Estimated effect of reopening on daily deaths per million

| week | state | treated-synthetic | std | Week | state | treated-synthetic | std |
| --- | --- | --- | --- | --- | --- | --- | --- |
| 0 | Colorado | -0.005 | 0.124 | 0 | Georgia | 0.103 | 0.171 |
| 1 | Colorado | -2.60*** | 0.433 | 1 | Georgia | 0.243 | 0.409 |
| 2 | Colorado | -4.5*** | 0.667 | 2 | Georgia | 2.081*** | 0.459 |
| 3 | Colorado | 18.2*** | 0.168 | 3 | Georgia | 4.222*** | 0.436 |
| 0 | Mississippi | 0.023 | 0.106 | 0 | Tennessee | -0.128** | 0.055 |
| 1 | Mississippi | 2.14*** | 0.186 | 1 | Tennessee | 2.05*** | 0.047 |
| 2 | Mississippi | 10.17*** | 0.213 | 2 | Tennessee | 1.593*** | 0.046 |
| 3 | Mississippi | 6.41*** | 0.208 | 3 | Tennessee | 2.085*** | 0.071 |
| 0 | Texas | 0.123 | 0.148 | 0 | Alabama | -0.035 | 0.095 |
| 1 | Texas | 5.12*** | 0.281 | 1 | Alabama | 3.182*** | 0.101 |
| 2 | Texas | 13.60** | 0.515 | 2 | Alabama | 8.232*** | 0.117 |
| 3 | Texas | 18.05*** | 0.474 | 3 | Alabama | 3.650*** | 0.108 |

Note: \*\*\* p<0.01, \*\* p<0.05, \* p<0.1. Standard errors are estimated by bootstrapping 50 times.

The results reported in Table A3 are a bit more volatile because of the relatively lower quality of the test data. But, the pattern is similar to what was reported in Table 5. Except for Georgia, the remaining 5 states that reopened before May 1^st^ did not increase their test capacity. This would have enabled identifying cases at an early stage and permitted contact tracing. Again, Texas saw the largest decline in tests performed per million population.

**Table A3.** Estimated effect of reopening on daily tests per million population

| week | state | treated-synthetic | std | week | state | treated-synthetic | std |
| --- | --- | --- | --- | --- | --- | --- | --- |
| 0 | Colorado | 5.0 | 12.6 | 0 | Georgia | -7.2 | 12.6 |
| 1 | Colorado | -1639.1*** | 321.0 | 1 | Georgia | 10.6 | 70.8 |
| 2 | Colorado | -2599.6*** | 273.6 | 2 | Georgia | -595.8*** | 170.4 |
| 3 | Colorado | -3025.0*** | 254.4 | 3 | Georgia | 5525.7*** | 54.1 |
| 0 | Mississippi | 1.0 | 10.5 | 0 | Tennessee | -4.1 | 15.5 |
| 1 | Mississippi | -838.0*** | 44.6 | 1 | Tennessee | 734.9*** | 62.2 |
| 2 | Mississippi | -3020.2*** | 83.1 | 2 | Tennessee | 658.1*** | 51.7 |
| 3 | Mississippi | 118.9 | 112.8 | 3 | Tennessee | -1844.7*** | 61.8 |
| 0 | Texas | -3.6 | 7.3 | 0 | Alabama | 0.8 | 14.5 |
| 1 | Texas | -3131.1*** | 640.6 | 1 | Alabama | -579.0*** | 25.4 |
| 2 | Texas | -9381.7*** | 1217.9 | 2 | Alabama | -1746.2*** | 43.8 |
| 3 | Texas | -8554.1*** | 821.0 | 3 | Alabama | -2718.2*** | 67.5 |

Note: \*\*\* p<0.01, \*\* p<0.05, \* p<0.1. Standard errors are estimated by bootstrapping 50 times.

## Appendix 2

In this Appendix I restrict the control group to the states that reopened after May 10^th^. The results are reported in Tables A4-A6, using the same notation as in Tables 2-4.

Table A4 reports the estimated effect of reopening on daily confirmed cases, with the donor pool being restricted to the states that reopened after May 10^th^. The results are quite similar to the results in Table 2. Reopening the economy early has significant and large effect on the increase in daily confirmed cases.

**Table A4.** Estimated effect of reopening on daily confirmed cases

| open | state | treated-synthetic | std | open | state | treated-synthetic | std |
| --- | --- | --- | --- | --- | --- | --- | --- |
| 0 | Colorado | 0.28 | 1.6 | 0 | Georgia | 0.74 | 2.2 |
| 1 | Colorado | 77.1 | 49.1 | 1 | Georgia | 265.2*** | 44.5 |
| 2 | Colorado | 175.8** | 74.2 | 2 | Georgia | 163.3*** | 49.4 |
| 3 | Colorado | 122.2* | 64.7 | 3 | Georgia | 202.1*** | 50.8 |
| 0 | Mississippi | -0.62 | 2.2 | 0 | Tennessee | -0.05 | 2.7 |
| 1 | Mississippi | 30.3** | 15.4 | 1 | Tennessee | 151.8*** | 14.2 |
| 2 | Mississippi | 101.6*** | 19.6 | 2 | Tennessee | 59.1*** | 15.3 |
| 3 | Mississippi | 94.9*** | 16.0 | 3 | Tennessee | 24.90 | 15.7 |
| 0 | Texas | 0.27 | 2.0 | 0 | Alabama | 1.01 | 2.1 |
| 1 | Texas | 489.0*** | 64.8 | 1 | Alabama | 70.6*** | 5.7 |
| 2 | Texas | 754.2*** | 72.2 | 2 | Alabama | 126.6*** | 6.1 |
| 3 | Texas | 806.3*** | 71.9 | 3 | Alabama | 173.4*** | 6.3 |

Note: I restrict the control group to the states that reopened after May 10<sup>th</sup>. \*\*\* p<0.01, \*\* p<0.05, \* p<0.1. Standard errors are estimated by bootstrapping 50 times.

Table A5 reports the estimated effect of reopening on deaths, with the donor pool restricted to the states that reopened after May 10^th^. The results are quite similar to the results in Table 3, except that the pre-treatment period for Tennessee doesn’t match very well. Reopening the economy early significantly increases deaths, and the magnitude is large.

**Table A5.** Estimated effect of reopening on daily deaths

| week | State | treated-synthetic | std | week | state | treated-synthetic | std |
| --- | --- | --- | --- | --- | --- | --- | --- |
| 0 | Colorado | -0.002 | 0.13 | 0 | Georgia | 0.098 | 0.17 |
| 1 | Colorado | -2.63*** | 0.44 | 1 | Georgia | 0.30 | 0.57 |
| 2 | Colorado | -4.45*** | 0.67 | 2 | Georgia | 2.07*** | 0.47 |
| 3 | Colorado | 18.25*** | 0.18 | 3 | Georgia | 4.22*** | 0.44 |
| 0 | Mississippi | 0.026 | 0.11 | 0 | Tennessee | -0.13** | 0.05 |
| 1 | Mississippi | 2.16*** | 0.21 | 1 | Tennessee | 2.05*** | 0.05 |
| 2 | Mississippi | 10.18*** | 0.24 | 2 | Tennessee | 1.60*** | 0.05 |
| 3 | Mississippi | 6.42*** | 0.22 | 3 | Tennessee | 2.09*** | 0.07 |
| 0 | Texas | 0.12 | 0.15 | 0 | Alabama | -0.034 | 0.10 |
| 1 | Texas | 5.21*** | 0.29 | 1 | Alabama | 3.19*** | 0.11 |
| 2 | Texas | 13.61*** | 0.52 | 2 | Alabama | 8.24*** | 0.12 |
| 3 | Texas | 18.06*** | 0.48 | 3 | Alabama | 3.65*** | 0.11 |

Note: I restrict the control group to the states that reopened after May 10<sup>th</sup>. \*\*\* p<0.01, \*\* p<0.05, \* p<0.1. Standard errors are estimated by bootstrapping 50 times.

Table A6 reports the estimated effect of reopening on test conducted, with the donor pool being restricted to the states that reopened after May 10^th^. The results suggest that except for Georgia, the number of tests performed in other treated states greatly decrease compared with their counterparts after they reopened the economy.

**Table A6.** Estimated effect of reopening on daily tests

| week | state | treated-synthetic | std | week | state | treated-synthetic | std |
| --- | --- | --- | --- | --- | --- | --- | --- |
| 0 | Colorado | 0.84 | 7.3 | 0 | Georgia | -7.50 | 13.8 |
| 1 | Colorado | -1367*** | 378.3 | 1 | Georgia | -95.8 | 147.1 |
| 2 | Colorado | -2413*** | 325.7 | 2 | Georgia | -1063*** | 193.4 |
| 3 | Colorado | -2911*** | 325.2 | 3 | Georgia | 5293*** | 241.7 |
| 0 | Mississippi | 3.3 | 14.6 | 0 | Tennessee | -0.20 | 10.1 |
| 1 | Mississippi | -680*** | 36.9 | 1 | Tennessee | 775*** | 110.5 |
| 2 | Mississippi | -2588*** | 47.4 | 2 | Tennessee | 540*** | 82.6 |
| 3 | Mississippi | 626*** | 53.4 | 3 | Tennessee | -1740*** | 73.6 |
| 0 | Texas | -6.98 | 8.5 | 0 | Alabama | 3.95 | 12.8 |
| 1 | Texas | -2934*** | 1420.8 | 1 | Alabama | -545*** | 39.8 |
| 2 | Texas | -9198*** | 1997.6 | 2 | Alabama | -1911*** | 58.2 |
| 3 | Texas | -8165*** | 2519.1 | 3 | Alabama | -2699*** | 65.4 |

Note: I restrict the control group to the states that reopened after May 10<sup>th</sup>. \*\*\* p<0.01, \*\* p<0.05, \* p<0.1. Standard errors are estimated by bootstrapping 50 times.

## Appendix 3

In this Appendix I assign weights to synthetic control states based on the variable of interest (i.e., daily confirmed cases, COVID-19 related deaths, number of tests conducted) on each of 14 pre-treatment dates. The results are reported in Tables A7-A9, using the same notation as in Tables 2-4.

Table A7 reports the estimated effect of reopening on daily confirmed cases, with the pre-treatment periods being extended from 7 days to 14 days. I find similar results as in Table 2: reopening the state significantly increases the daily confirmed cases.

**Table A7.** Estimated effect of reopening on daily confirmed cases

| week | state | treated-synthetic | std | week | state | treated-synthetic | std |
| --- | --- | --- | --- | --- | --- | --- | --- |
| 0 | Colorado | 2.11 | 1.34 | 0 | Georgia | -0.49 | 2.06 |
| 1 | Colorado | 150.6*** | 38.45 | 1 | Georgia | 240.5*** | 48.80 |
| 2 | Colorado | 195.2*** | 54.50 | 2 | Georgia | 156.0*** | 52.90 |
| 3 | Colorado | 135.1*** | 54.81 | 3 | Georgia | 213.9*** | 52.81 |
| 0 | Mississippi | -0.45 | 2.57 | 0 | Tennessee | -0.56 | 2.65 |
| 1 | Mississippi | 44.1*** | 8.56 | 1 | Tennessee | 188.0*** | 8.11 |
| 2 | Mississippi | 111.5*** | 15.31 | 2 | Tennessee | 90.5*** | 9.22 |
| 3 | Mississippi | 109.0*** | 12.55 | 3 | Tennessee | 55.4*** | 9.17 |
| 0 | Texas | -0.32 | 1.62 | 0 | Alabama | 0.08 | 1.90 |
| 1 | Texas | 545.4*** | 100.47 | 1 | Alabama | 57.6*** | 7.90 |
| 2 | Texas | 792.5*** | 103.97 | 2 | Alabama | 116.3*** | 7.47 |
| 3 | Texas | 845.5*** | 101.86 | 3 | Alabama | 163.4*** | 6.71 |

Note: \*\*\* $p < 0.01$ , \*\* $p < 0.05$ , \* $p < 0.1$ . Standard errors are estimated by bootstrapping 50 times.

Table A8 reports the estimated effect of reopening on deaths, with the pre-treatment periods being extended from 7 days to 14 days. The results are quite similar to the results in Table 3, except that the pre-treatment period for Tennessee doesn’t match very well. Reopening the economy early significantly increases deaths, and the magnitude is large.

**Table A8.** Estimated effect of reopening on daily deaths

| week | state | treated-synthetic | std | week | state | treated-synthetic | std |
| --- | --- | --- | --- | --- | --- | --- | --- |
| 0 | Colorado | 0.039 | 0.13 | 0 | Georgia | -0.075 | 0.18 |
| 1 | Colorado | 4.49*** | 0.67 | 1 | Georgia | 0.48 | 0.43 |
| 2 | Colorado | 2.20*** | 0.18 | 2 | Georgia | 2.17*** | 0.54 |
| 3 | Colorado | 21.18*** | 0.18 | 3 | Georgia | 4.25*** | 0.50 |
| 0 | Mississippi | 0.19 | 0.15 | 0 | Tennessee | -0.16*** | 0.04 |
| 1 | Mississippi | 2.44*** | 0.25 | 1 | Tennessee | 1.46*** | 0.05 |
| 2 | Mississippi | 10.42*** | 0.26 | 2 | Tennessee | 1.20*** | 0.06 |
| 3 | Mississippi | 6.56*** | 0.24 | 3 | Tennessee | 1.90*** | 0.08 |
| 0 | Texas | 0.02 | 0.17 | 0 | Alabama | -0.053 | 0.10 |
| 1 | Texas | 8.03*** | 0.31 | 1 | Alabama | 2.12*** | 0.13 |
| 2 | Texas | 15.20*** | 0.53 | 2 | Alabama | 7.36*** | 0.16 |
| 3 | Texas | 19.22*** | 0.48 | 3 | Alabama | 2.91*** | 0.15 |

Note: \*\*\* p<0.01, \*\* p<0.05, \* p<0.1. Standard errors are estimated by bootstrapping 50 times.

Table A9 reports the estimated effect of reopening on test conducted, with the pre-treatment periods being extended from 7 days to 14 days. The results suggest that except for Georgia and Tennessee, the number of tests performed in other treated states greatly decrease compared with their counterparts after they reopened the economy.

**Table A9.**
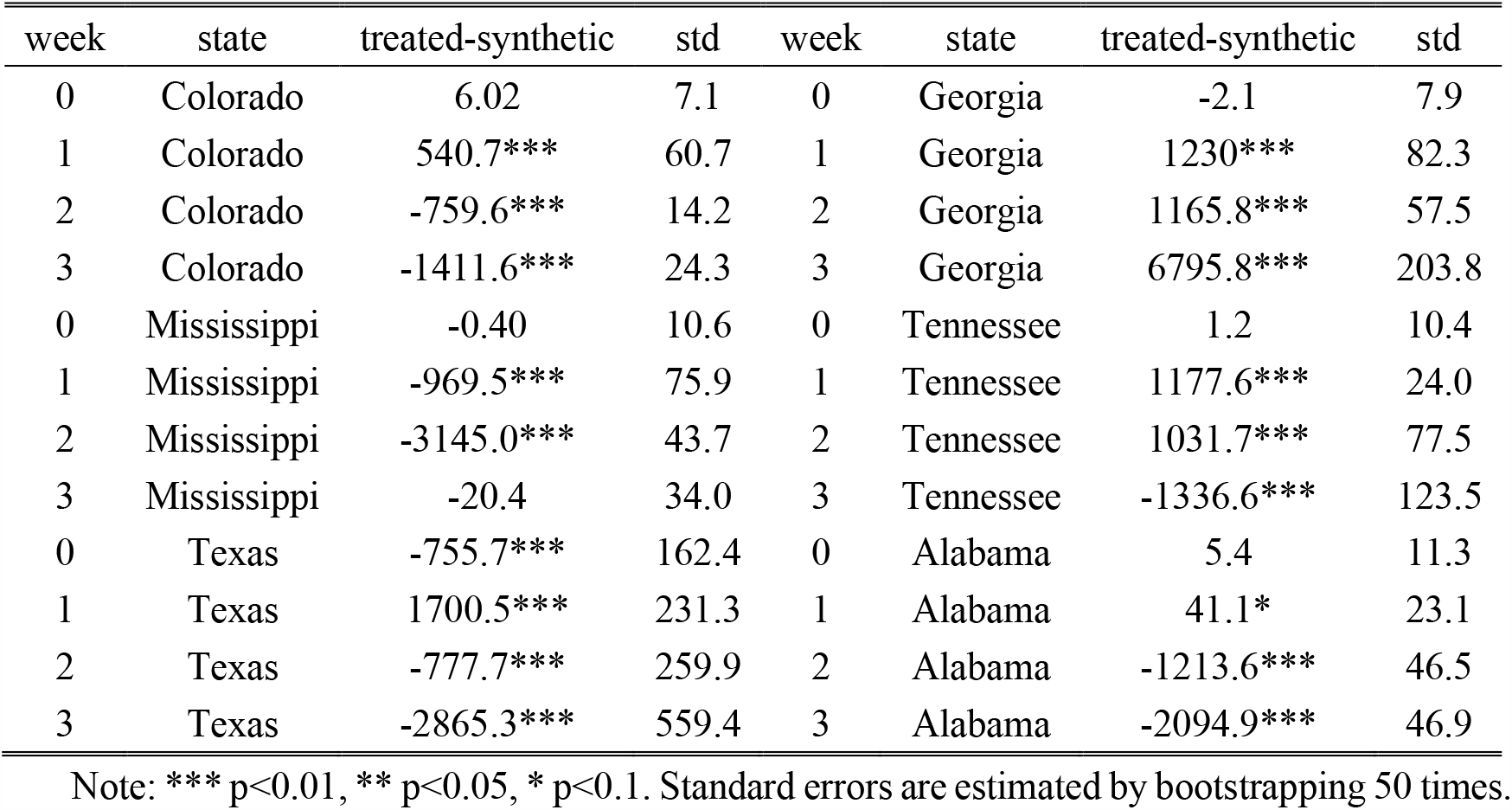
Estimated effect of reopening on daily tests

## Footnotes

2 https://covidtracking.com/api

3 https://www.kff.org/coronavirus-policy-watch/lifting-social-distancing-measures-in-america-state-actions-metrics/

4 Colorado, Delaware, Georgia, Maine, Mississippi, Missouri, New Hampshire, Pennsylvania, Texas, Vermont and Virginia mix the viral test and antibody test data.

5 The average number of daily tests performed at week 0 is 5,684.

## Notes

### Competing Interest Statement

The authors have declared no competing interest.

### Clinical Trial

This study doesn't involve any clinical trial. I use the publicly available data to assess the social and economic impact of reopening the economy in the US during COVID-19 pandemic.

### Funding Statement

No external funding was received

